# Distinct Metabolic Signatures Distinguish Lung, Colorectal and Ovarian Cancer

**DOI:** 10.64898/2026.04.08.26350309

**Authors:** Ioanna Tsiara, Efstathia Vouzaxaki, Joakim Ekström, Natallia Rameika, Fan Yang, Abhishek Jain, Alberto Iglesias Alonso, Tobias Sjöblom, Daniel Globisch

## Abstract

Cancer-related casualties are the most common cause of death worldwide. The discovery of biomarkers is of utmost importance for diagnosis and disease monitoring. Herein, we performed a comprehensive metabolomics biomarker discovery effort in plasma from 615 lung, ovarian and colorectal cancer patients at diagnosis and 95 non-cancerous control subjects. This pan-cancer investigation identified specific panels of metabolites in the entire sample cohort with a high discriminating power and demonstrated by combined ROC AUC values of up to 0.95. The identified metabolites are mainly associated with lipid and amino acid metabolism as well as xenobiotic transformation. These metabolite panels of high predictive power provide new metabolic insights in these cancers and demonstrate the potential of metabolomics for improved diagnosis and monitoring disease progression.

## Introduction

Cancer represents a significant global health issue and remains the leading cause of morbidity and mortality worldwide ^1^. Lung, colorectal, and ovarian cancers collectively account for 30% of global cancer-related deaths (18.7%, 9.3% and 2.1% respectively) ^2^. Lung cancer is the leading cause of cancer death worldwide, representing the first and third leading cause of death in men and women, respectively, accounting for 12% of all diagnosed cancers with 2.5 million new cases in 2022 ^1–4^. Lung cancer continues to have poor outcomes, particularly when diagnosed at advanced stages. Although five-year survival rates for all types of lung and bronchus cancer are 28.1% (2015-2021), when the disease is detected and treated at an early stage, this rate can increase to 64.7%. One of the primary reasons for such a high degree of severity of lung cancer is the low rates of early diagnosis, associated primarily with the absence of clear clinical symptoms at an initial stage. This is further challenged by the lack of effective screening programs for this cancer type. Similarly, colorectal cancer (CRC) ranks as the third most common cancer globally and is the second cause of cancer-related mortality ^3–5^. Early diagnosis is crucial for survival, as the 5-year survival rate for patients diagnosed with stage I reaches up to 90% ^6^. In contrast, this rate drops to ∼12% in cases where distant metastases are present at diagnosis ^7,8^. Ovarian cancer is the seventh most frequently diagnosed cancer among women globally and represents the most lethal gynecological cancer, mainly attributed to limitations of detecting the disease at an early stage ^9^. This diagnostic difficulty stems from the asymptomatic nature of the disease at an early stage, with nearly 60% of patients having already developed metastases at the time of diagnosis ^10,11^. Although mortality varies across the world, it is generally proportional to disease stage at the time of diagnosis ^12^. At stage I, the 5-year survival is 92%, but at FIGO stage III–IV, the 5-year survival rate is only 29% ^13,14^. In addition to the high mortality rate resulting from late diagnosis, the lack of specific treatment options for ovarian cancer makes classification, diagnosis, and treatment difficult ^15,16^. Survival outcomes are significantly improved when complete cytoreductive surgery can be achieved, as the absence of residual disease after surgery, even in advanced-stage ovarian cancer, has been associated with markedly prolonged survival ^17^.

Even with advancements in diagnosis and treatment options, more precise methods are required for early detection, prognosis, and monitoring of lung, colorectal, and ovarian cancers ^18^. In recent years, metabolomics has emerged as a powerful tool for the investigation of dynamic biochemical processes. Especially plasma metabolomic profiling has been increasingly applied to investigate diverse diseases individually including lung, colorectal, and ovarian cancer, which have revealed systematic metabolic alterations associated with cancer development and progression ^18,19^.These studies have consistently identified fatty acids and lipids, including lysophosphatidylethanolamines (LysoPEs), phosphatidylcholines (PCs) and sphingolipids ^20^, and small organic acids (β-hydroxybutyrate) ^21^, crucial metabolic intermediates in glycolysis and the Calvin cycle ^22^ as recurrent biomarkers for lung cancer ^23–26^. In colorectal cancer, the main findings include alterations of amino acids ^27^, nucleotide metabolites such as uridine, and pyrimidine metabolism ^28^. Reports of acid-type biomarkers were published for ovarian cancer ^29,30^ such as tryptophan pathway metabolites, lactate and TCA metabolism, and carnitine metabolites ^29,31^. However, the validation of biomarkers is missing in most cases, while another limitation is the lack of identification of cancer type-specific metabolites to identify overlapping metabolic signatures that can be excluded for diagnostics. By identifying these biochemical processes that drive cellular phenotypes, metabolomic profiling opens the possibility to monitor treatment efficacy by capturing metabolic alterations in response to chemotherapy or other interventions, supporting real-time adjustments of clinical interventions ^5^.

While the complexity of metabolomic data represents meaningful biological insights, it requires the application of advanced biostatistical and computational tools for accurate interpretation ^32^. The prevalence and survival statistics across lung, colorectal, and ovarian cancers underscore the urgent need for the discovery of cancer type-specific biomarkers that could facilitate early detection and support personalized treatment strategies. In this study, an untargeted metabolomics approach was applied to analyse plasma samples from patients at four different stages of lung, colorectal, and ovarian cancers. We identified a series of metabolites from different metabolic pathways that were consistently altered among the cancers and within cancer stages. Using a receiver operating characteristic (ROC)-based biomarker discovery approach, a distinct pattern of alterations was observed in metabolite panels across different types of cancer. Furthermore, this study assessed the clinical relevance of specific metabolites identified as potential biomarkers, aiming to both validate these findings within these current patient cohorts and to contribute to a broader understanding of metabolism in cancer diagnosis ^33,34^.

## Results

### Cohort characteristics and metabolomics workflow

The investigated cohort consisted of 710 plasma samples derived from patients enrolled in the U-CAN biobank (**Figure 1A**) ^35^. From this biobank, 615 patients were diagnosed with lung (LuCa), colorectal (CRC), or ovarian cancer (OvCa) across stages I–IV. An additional 95 patients with non-malignant diagnoses for the respective cancer types were included as non-cancer controls (Ctrls) and used as a reference group. For a robust biomarker discovery pipeline, our cohort was divided into three independent analytical batches, where samples were randomized, ensuring a balanced and unbiased distribution of cancer cases and controls across each investigated dataset (**Figure 1A / Table 1**). The batches were assigned as Batch A (n = 229), Batch B (n = 239) and Batch C (n =242). The sex distribution across the full cohort was 64.6% female in Batch A, 59.4% in B, and 64.5% in C. The mean ages were 65.4 (IQR: 9.0), 65.6 (IQR: 11.5), and 63.5 (IQR: 12.0) years, respectively (**Figures 1B-C**). Untargeted mass spectrometry-based metabolomics analysis was performed separately for each batch in positive and negative ionization modes. To account for potential batch effects and technical variations over the LC–MS sequence of each batch, we applied Systematic Error Removal using Random Forest (SERRF), a QC-based normalization tool in R (**Figure S1**).^36^

**Figure 1.**
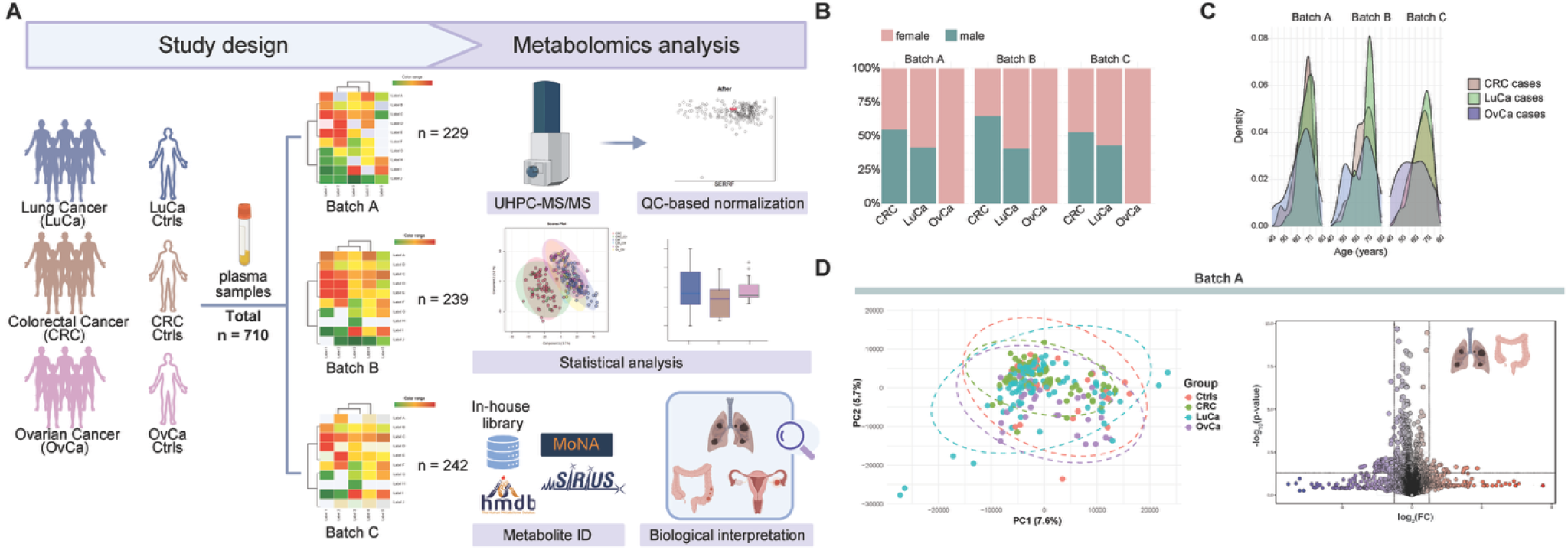
Overview of the study design and cohort characteristics. A) The investigated study cohort consisting of patients with lung, colorectal or ovarian cancer. The cohort was divided in three independent batches, and our standard metabolomics workflow was applied. B) Percentage of female and male cases by cancer type and batch. C) Age density plot for the cancer cases per batch. D) PCA scores plot of all groups and volcano plot (*p* < 0.05, F.C = 2.0) of the CRC vs LuCa groups for Batch A. Statistical comparisons between groups were performed using one-way ANOVA with Tukey’s HSD post-hoc analysis.

**Table 1.** Demographic and clinical characteristics of cancer cases and controls by batch. The percentage of non-cancer controls (ctrls) was calculated based on the whole cohort, whereas the percentage indicated in each cancer stage was based on the cancer group cases only.

| Batch | Group | n | Mean Age | Female | Stage I | Stage II | Stage III | Stage IV |
| --- | --- | --- | --- | --- | --- | --- | --- | --- |
| Batch A | Ctrl LuCa | 6 (6.3%) | 65 (8.0) | 4 (67%) | - | - | - | - |
|  | Ctrl CRC | 10 (12%) | 67.8 (5.1) | 8 (80%) | - | - | - | - |
|  | Ctrl OvCa | 15 (29%) | 61.1 (9.7) | 15 (100%) | - | - | - | - |
|  | LuCa cases | 89 | 67.4 (5.5) | 52 (58%) | 40 (45%) | 10 (11%) | 12 (14%) | 27 (30%) |
|  | CRC cases | 73 | 64.7 (6.6) | 33 (45%) | 8 (11%) | 10 (14%) | 36 (49%) | 19 (26%) |
|  | OvCa cases | 36 | 63.1 (10) | 36 (100%) | 13 (36%) | 5 (14%) | 12 (33%) | 6 (17%) |
| Batch B | Ctrl LuCa | 7 (6.8%) | 61.4 (8.9) | 5 (71%) | - | - | - | - |
|  | Ctrl CRC | 14 (17%) | 69.4 (3.9) | 4 (29%) | - | - | - | - |
|  | Ctrl OvCa | 13 (26%) | 57.4 (8.9) | 13 (100%) | - | - | - | - |
|  | LuCa cases | 96 | 67.3 (6.6) | 57 (59%) | 47 (49%) | 8 (8%) | 17 (18%) | 24 (25%) |
|  | CRC cases | 71 | 65.4 (7.2) | 25 (35%) | 5 (7%) | 22 (31%) | 33 (47%) | 11 (16%) |
|  | OvCa cases | 38 | 63.7 (9.7) | 38 (100%) | 10 (26%) | 2 (5%) | 15 (40%) | 11 (29%) |
| Batch C | Ctrl LuCa | 4 (3.9%) | 55.2 (11.5) | 2 (50%) | - | - | - | - |
|  | Ctrl CRC | 13 (15%) | 65.5 (7.5) | 9 (69%) | - | - | - | - |
|  | Ctrl OvCa | 13 (24%) | 59.5 (10.7) | 13 (100%) | - | - | - | - |
|  | LuCa cases | 98 | 65.3 (6.9) | 56 (57%) | 35 (36%) | 10 (10%) | 23 (24%) | 30 (31%) |
|  | CRC cases | 72 | 64.5 (7.6) | 34 (47%) | 6 (8%) | 23 (32%) | 31 (43%) | 12 (17%) |
|  | OvCa cases | 42 | 59.1 (11.1) | 42 (100%) | 8 (19%) | 3 (7%) | 13 (31%) | 18 (43%) |
| Total | Ctrl LuCa | 17 (18%) | 60.5 | 11 | - | - | - | - |
|  | Ctrl CRC | 37 (39%) | 67.6 | 21 | - | - | - | - |
|  | Ctrl OvCa | 41 (43%) | 59.3 | 41 | - | - | - | - |
|  | LuCa cases | 283 | 66.7 | 165 | 122 (43%) | 28 (10%) | 52 (18%) | 81 (29%) |
|  | CRC cases | 216 | 64.9 | 92 | 19 (9%) | 55 (26%) | 100 (46%) | 42 (19%) |
|  | OvCa cases | 116 | 62 | 116 | 31 (27%) | 10 (9%) | 40 (34%) | 35 (30%) |

To facilitate discovery of biomarkers for detection of early-stage cancers, we grouped the patients by early (stage I/II) and late (stage III/IV) stage tumors. This allowed us to investigate metabolic alterations associated with disease progression while maintaining sufficient sample size per cancer stage group. Additionally, we performed principal component analysis (PCA) to reduce data dimensionality and to identify patterns across all cancer types (**Figure 1D** / left). The PCA analysis did not reveal clear separation between the three cancer groups and controls, suggesting that these cancers may share overlapping metabolic profiles at a global metabolic level. With a focus on investigating the metabolic differences between LuCa and CRC that occur in the total population, univariate analysis (t-test, *p* < 0.05) identified significant alterations of 608 unique features in Batch A, 404 in Batch B and 943 in Batch C in the positive ionization mode (**Figures 1D** / right).

### Shared and cancer-specific metabolic signatures

First, we sought to identify metabolites significantly altered between early-stage (I/II) and late-stage (III/IV) among all three cancer types. For this statistical analysis, OvCa was only investigated within its stages (I/II vs. III/IV). We considered metabolites that showed consistent changes in at least two out of the three independent batches. Notably, 3 metabolites were identified to be consistently altered in all 3 batches. 3-Hydroxypyridine sulfate was found to be upregulated in stage I/II of LuCa compared to stage III/IV of CRC (**Figure 2A**). The same trend was observed for *S-*prenyl-L-cysteine, which was increased in the early-stage LuCa compared with early-stage CRC, in two batches (**Figure 2B**). One lysophospholipid, LysoPC(20:3), showed decreased abundance in late stages of LuCa compared to CRC (**Figure 2C**). In Batch A and C, this dysregulation was also observed towards stage I/II of CRC.

**Figure 2.**
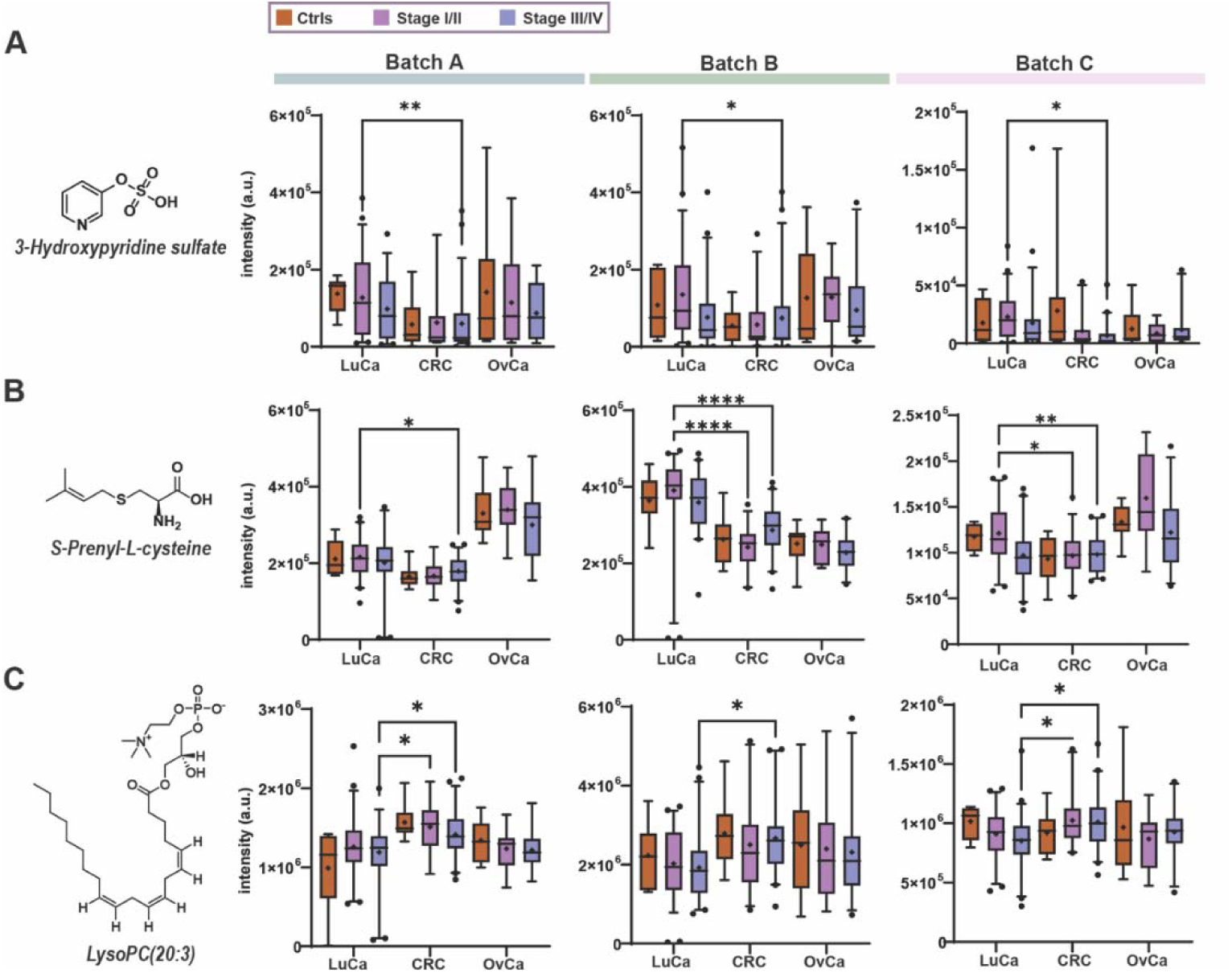
Overview of three metabolites consistently altered in all three batches. Univariate analysis of the statistically significant metabolites across all three independent batches (one-way ANOVA with Tukey’s HSD post-hoc analysis, ****p ≤ 0.0001; **p ≤ 0.01; *p ≤ 0.05). **A)** 3-Hydroxypyridine sulfate, **B)** *S*-prenyl-L-cysteine and **C)** lysoPC(20:3).

We also identified a series of metabolites that had concordant and significant differences in two of the batches, and concordant but non-significant differences in the remaining batch. However, due to the consistent direction of change in all three batches, these compounds may still have potential utility in distinguishing disease and disease stage. Benzoic acid was downregulated in LuCa stages I/II and III/IV in comparison to CRC (**Figures 3A, S2A**). The metabolite *S*-propenylcysteine displayed the opposite trend and was significantly upregulated in stage I/II of LuCa compared to all stages of CRC (**Figures 3B, S2B**). Phenol sulfate was found to differentiate late-stage CRC and LuCa, with decreased intensities in the latter, which was consistent for Batch A and C. In addition, it was observed as a strong trend in Batch B (fold CRC/LuCa: 2.01 / **Figures 3C, S2C**).

**Figure 3.**
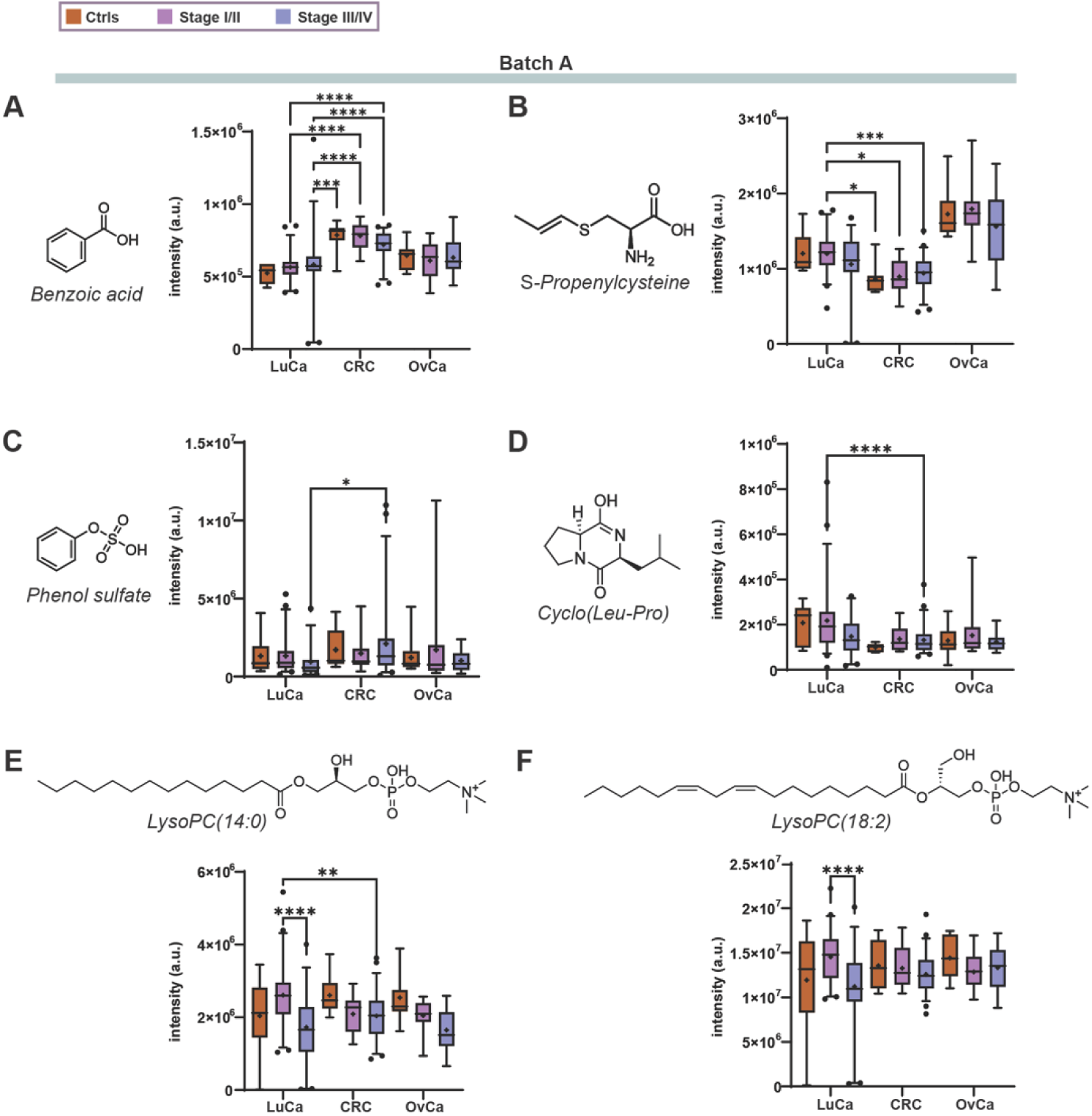
Overview of six statistically altered cancer-type and stage-specific metabolites. Univariate analysis of the statistically significant metabolites in two independent batches (one-way ANOVA with Tukey’s HSD post-hoc analysis, \*\*\*\**p* ≤ 0.0001; \*\*\**p* ≤ 0.001; \*\**p* ≤ 0.01; \**p* ≤ 0.05). Representative example of Batch A for **A)** benzoic acid, **B)** *S*-propenylcysteine, **C)** phenol sulfate, **D)** cyclo(Leu-Pro), **E)** lysoPC (14:0), and **F)** lysoPC (18:2).

The dipeptide cyclo(Leu-Pro) was also altered between CRC and LuCa and was downregulated in late-stage CRC compared to early-stage LuCa (**Figures 3D, S2D**). Lastly, two glycerophospholipids, lysoPC(14:0) and lysoPC(18:2), were identified to be significantly downregulated with LuCa disease progression (**Figures 3E-F, S2E**-**F**). Additionally, lysoPC(14:0) was altered between stage I/II of LuCa and stage III/IV of CRC, with decreased levels observed in CRC.

### Sex-dependent investigation across the three cancer types

We next searched for metabolic differences across the three cancer types, solely focusing our analysis on female patients. Initially, the PCA analysis did not reveal any distinct separation between cancer types. Instead, the pan-cancer datasets demonstrated substantial overlap in two-dimensional PCA space, suggesting shared metabolic profiles across the cancer types in female patients (**Figure 4A**). For the pan-cancer analysis, we combined all stages (I-IV) for each cancer type and all non-cancer controls were grouped together. We then compared the three cancers using one-way ANOVA with Tukey’s HSD post-hoc analysis, to identify cancer-specific metabolic changes. Among our findings, benzoic acid was found to be significantly upregulated in CRC compared to both LuCa and OvCa in two independent batches (**Figure 4B**). In Batch C, where it was not altered, the increase of the metabolite in CRC was 4.0% and 6.6% compared to LuCa and OvCa, respectively (**Figure S3A**). Another metabolite that was consistently altered in two out of three datasets was sphingosine, exhibiting upregulation in LuCa and CRC compared to the non-cancer controls, and consistent normal levels in OvCa (**Figures 4C, S3B**). Lastly, the two glycerophospholipids that were found significantly altered in the total population cohort, lysoPC(14:0) and lysoPC(18:2), remained consistently dysregulated also in the sex-specific analysis, underscoring their potential as putative pan-cancer biomarkers.

**Figure 4.**
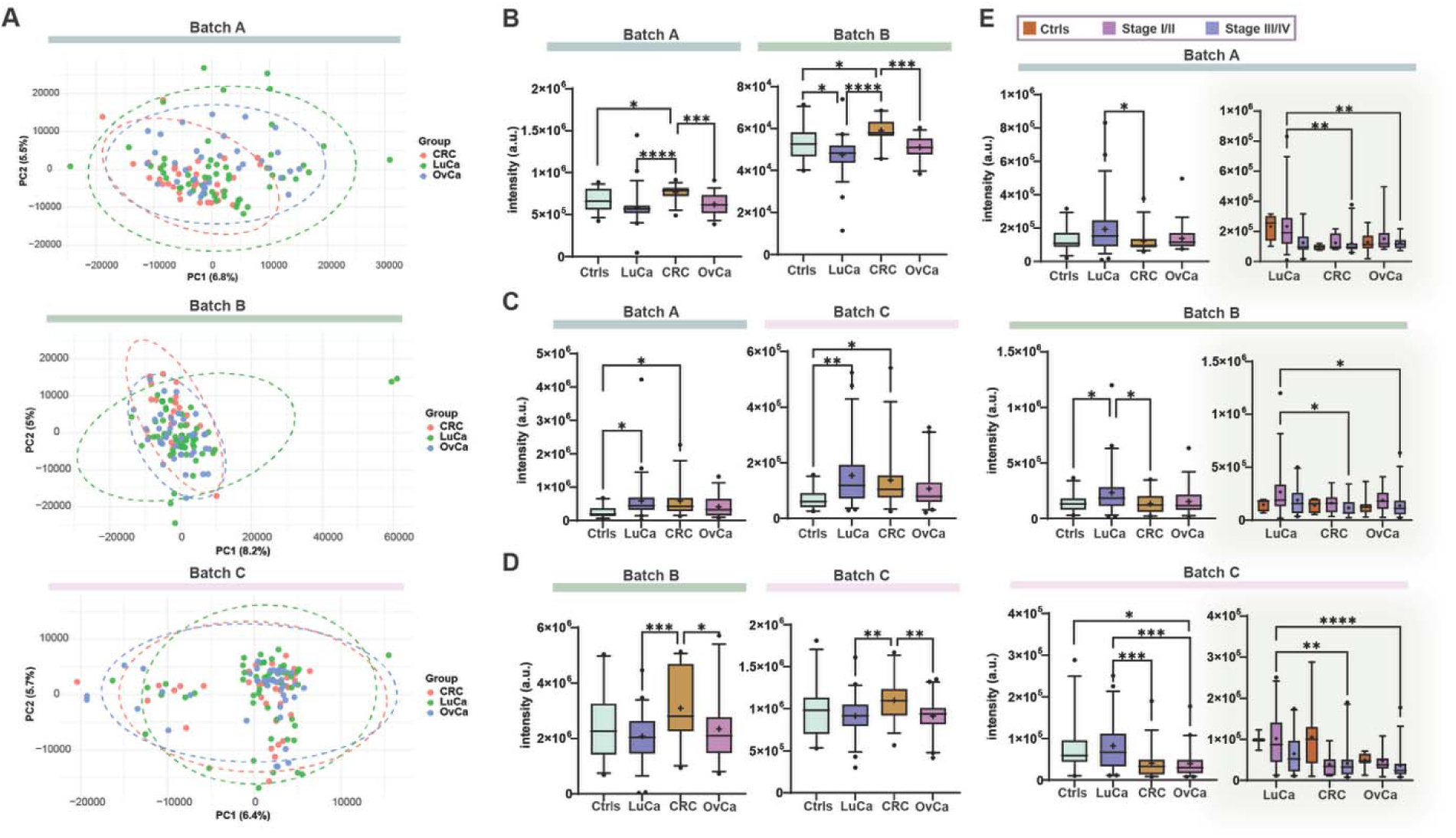
Pan-cancer sex-dependent investigation of female individuals. A) PCA scores plots for the pan-cancer analysis of female individuals in the three batches. Representative batches for B) benzoic acid, C) sphingosine, D) lysoPC(20:3), and E) cyclo(Leu-Pro) with additional stage-dependent analysis in all three batches.

Two metabolites showed consistent alteration in all three batches. LysoPC(20:3) showed an increase in CRC compared to OvCa, and interestingly, the same trend was observed compared to LuCa, in 2/3 batches (except Batch A, fold CRC/LuCa: 1.1/ **Figures 4D, S3C**). The second metabolite that was consistent among batches was cyclo(Leu-Pro) that was downregulated in CRC compared to LuCa, similarly as in the total cohort analysis (**Figure 4E**). We next investigated the alterations of this metabolite in the different cancer stages. Beyond the expected dysregulation between early-stage LuCa and late-stage CRC, we observed upregulation of stage I/II of LuCa towards stage III/IV of OvCa.

The investigation of the male population across LuCa and CRC with univariate analysis (p < 0.05, F.C. = 2.0) revealed 205 features upregulated and 80 downregulated in CRC in Batch A, 148 features upregulated and 50 downregulated in CRC in Batch B and 58 features upregulated and 46 downregulated in CRC in Batch C (**Figure S4**). Among the batches, 23 unique significantly altered features were common. The majority of these compounds was either of low intensity or were identified as drugs and other exogenous molecules. These findings suggest that these compounds are not ideal as metabolite biomarkers for disease diagnostics, since they are not of an endogenous nature.

### Candidate biomarker discovery

As the next step, we investigated potential metabolic biomarkers to distinguish between LuCa and CRC in both sexes combined as well as differences between the individual cancer stages. For this purpose, we combined all analytical batches and applied ComBat normalization to minimize batch effects and improve sample comparability before building classification models for ROC analysis (**Figure S5**) ^37^. After merging the batches, we did not observe any separation trends for the cancer groups in the PCA analysis in line with the analysis *vide supra* of each individual batch (**Figure S6**).

In the comparison of early-stage cases (stage I/II LuCa vs. CRC), a panel of nine metabolites achieved a combined AUC of 0.95, indicating excellent discriminatory power (**Figure 5A**). The top-performing metabolites were *S*-prenyl-L-cysteine (AUC: 0.82), 2-furoylglycine (AUC: 0.67), 3-hydroxypyridine sulfate (AUC: 0.71), lysoPE(18:3) (AUC: 0.64), 2,5-furandicarboxylic acid (AUC: 0.63), dodecanoyl-sn-glycero-3-phosphocholine (AUC: 0.62), octadecylamine (AUC: 0.59), 4-hydroxybenzoic acid (AUC: 0.86), and *S*-propenylcysteine (AUC: 0.82). This metabolite panel contributes to a highly discriminative signature between the two cancer types in early development (**Figure S7A**).

**Figure 5.**
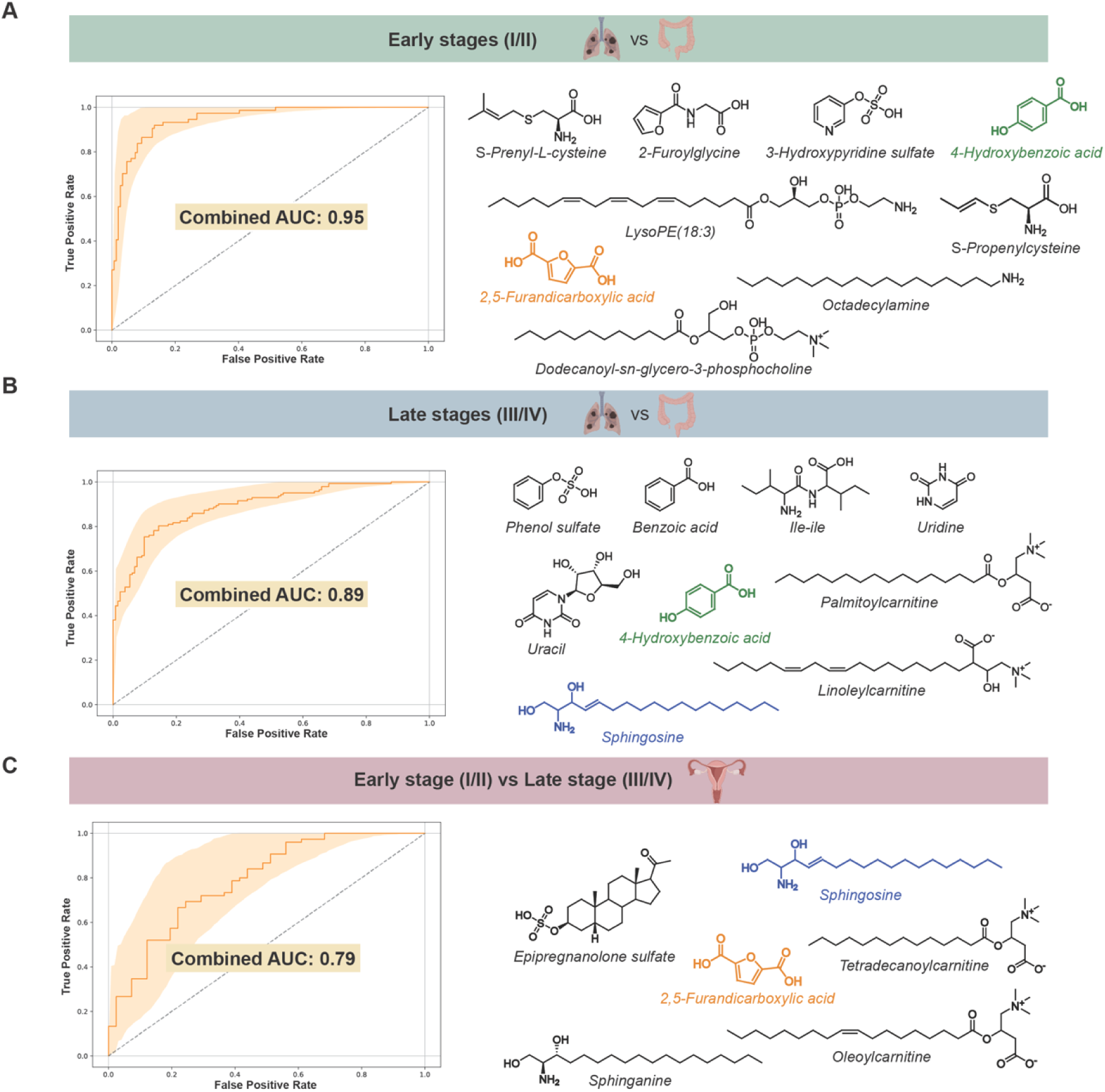
ROC curve analysis for the different candidate biomarker discovery analyses. Combined ROC curve with chemical structures of the top candidates for **A)** early-stage (I/II) LuCa vs CRC, **B)** late-stage (III/IV) LuCa vs CRC and **C)** early-stage vs late-stage OvCa. The coloured metabolites were identified in at least two comparisons. The highlighted area in the ROC curves represent the 95% confidence interval.

The performance of the ROC model remained strong when comparing late-stage (stage III/IV) LuCa and CRC, achieving a combined AUC of 0.89 for a panel of nine metabolites (**Figure 5B, S7B**). This result also reveals a high level of accuracy and highlights the utility of metabolomics for differentiating advanced cancer stages. The key metabolites driving this classification included phenol sulfate (AUC: 0.72), benzoic acid (AUC: 0.69), Ile-Ile (AUC: 0.69), uridine (AUC: 0.68), palmitoylcarnitine (AUC: 0.63), 4-hydroxybenzoic acid (AUC: 0.85), linoleylcarnitine (AUC: 0.70), uracil (AUC: 0.67), and sphingosine (AUC: 0.66). Notably, most of these compounds differ from those observed in the early-stage discrimination, suggesting metabolic changes dependent on disease progression (**Figure S6**).

Interestingly, our results demonstrate that a metabolite panel can also differentiate between early and late stages of OvCa. The discriminatory power of the selected metabolites was lower, however, indicated good predictive ability based on a combined AUC of 0.79 for a panel of six metabolites (**Figure 5C**). Identifying metabolite trends associated with disease progression is an important step to improve prognostic assessments and clinical monitoring. The metabolites with the highest discriminatory power in this comparison were epipregnanolone sulfate (AUC: 0.71), 2,5-furandicarboxylic acid (AUC: 0.66), sphinganine (AUC: 0.69), sphingosine (AUC: 0.70), tetradecanoylcarnitine (AUC: 0.61), and oleoylcarnitine (AUC: 0.58) (**Figure S8A**). The CRC-specific model achieved a combined AUC of 0.69, while the LuCa-specific model yielded a combined AUC of 0.77 (**Figure S8B**-**C**).

## Discussion

In this study, we identified a series of metabolites that differ significantly across the three cancer types, colorectal, lung, and ovarian cancer. We have performed a comprehensive metabolomics analysis of well-characterized human plasma sample cohorts that identified key metabolic alterations and metabolite panels that represent candidate biomarkers for diverse clinically relevant comparisons for these three cancer types.

The three most promising metabolites are 3□hydroxypyridine sulfate, *S*-prenyl-L-cysteine and lysoPC(20:3) that were consistently identified in our analysis of three independent and randomly prepared cohort subset samples for comparison of early (I/II) and late (III/IV) cancer stages. One of the metabolites that was significantly increased in LuCa (stages I/II) compared to CRC (stages III/IV) is 3□hydroxypyridine sulfate. This metabolite is a xenobiotic conjugate and has been inversely associated with a risk for gastric cancer, especially in women and non□smokers ^38^. Interestingly, the compound class of hydroxypyridines has also been associated with bacterial metabolism with the precursor 3*-*hydroxypyridine as a product of bacterial biosynthesis ^39^. The second metabolite *S*-prenyl-L-cysteine was identified as a candidate marker upregulated at an early stage [LuCa (I/II) > CRC (I/II)]. This sulfur-containing amino acid produced from protein prenylation is considered a crucial post-translational modification for protein function. While cysteine metabolism is heavily involved in cancer through processes like redox balance and glutathione modifications, studies on human cancer focus on the broader biology of cysteine rather than the measurement of *S*-prenyl-L-cysteine specifically. The third consistently altered metabolite is lysoPC(20:3) that was identified to be reduced in LuCa (III/IV) compared to CRC (III/IV). LysoPCs are membrane-associated lipids with pro-inflammatory functions and in previous studies have been found upregulated in the plasma of LuCa patients ^40^.

Other metabolites that were altered in two out of three independent batches may hold potentially high biological value. One altered metabolite is benzoic acid that has previously been negatively correlated with CRC stage in serum and displayed strong cytotoxicity against LuCa and CRC cell lines ^41,42^. This metabolite was also dysregulated in the female cohorts, suggesting no bias based on sex. The metabolite *S*-propenylcysteine, that was found altered between LuCa and CRC, is known to modulate inflammatory and stress-related signaling pathways (MyD88 and NF-κB-dependent signaling). These properties suggest that altered *S*-propenylcysteine levels may reflect inflammation-associated metabolic reprogramming and changes in sulfur amino acid metabolism within the tumor microenvironment ^43,44^. Phenol sulfate that was identified to be reduced in LuCa (III/IV) compared to CRC (III/IV) is produced primarily in the liver during phase II metabolism and has been investigated in relation to LuCa and CRC before ^45^. Additionally, genetic variation in the phenol sulfotransferase has been linked with CRC. The origin of the metabolite is through microbiome metabolism to obtain phenol, which is afterwards sulfated by the human clearance mechanism. From the selective investigation of female patients due to the OvCa cases, we also have observed sphingosine to be altered, which is a key lipid in cell membranes and signaling that is central to sphingolipid metabolism. Disruptions in this pathway have been linked to a higher risk to develop lung cancer ^46,47^.

Cyclo(Leu-Pro) is a cyclic dipeptide reported to reduce cell migration and growth in aggressive breast cancer cells ^48^, which we found altered between CRC vs. LuCa and LuCa (I/II) vs. OvCa (III/IV) in the female population as well as in the total cohort. Disruptions in this pathway have been linked to a higher risk of developing lung cancer ^46,47^. Lastly, two glycerophospholipids were found significantly upregulated in LuCa (I/II) in the total population cohort, LysoPC(14:0) and LysoPC(18:2). These two LysoPCs and Cyclo(Leu-Pro) also remained consistently dysregulated in the sex-specific analysis, underscoring their potential as putative pan-cancer biomarkers. This observation is interesting and is in line with previous reports as specific glycerophospholipid species, including lysoPE(18:3), lysoPC(14:0) and lysoPC(18:3), have been reported to discriminate between LuCa patients and healthy individuals ^49,50^. In plasma, lysoPC(20:3) and lysoPC(18:2) have been found significantly elevated in cancer cases, with lysoPC(20:3) reported as a strong marker to distinguish stage I non-small cell lung cancer (NSCLC) from healthy controls ^51^. Interestingly, this elevation was evident in stage I/II disease but not in stage III/IV NSCLC that suggests a stage-dependent role. This metabolite was also consistently altered in all three independent batches in the female population between CRC and OvCa. Furthermore, LysoPC(14:0) has shown high discriminative power in differentiating drug-induced lung disease (DILD) from other lung conditions and appears associated with disease severity ^52^.

To determine the diagnostic value of these metabolites and identify additional key molecules, we performed ROC curve analysis, for which we combined the three independent batch analyses. The comparison of LuCa vs. CRC at an early stage, with an excellent AUC value of 0.95, revealed several metabolites that are strongly influenced by the diet, the environment or the microbiome metabolism in addition to any cancer-related changes. The two furan-based metabolites, 2□Furoylglycine and 2,5□furandicarboxylic acid, are mainly derived from diet or are produced by the microbiome and may act as indicators for these exposures ^53,54^. The already identified metabolite *S*-propenylcysteine in the univariate analysis that is an active compound with reported immunomodulatory effects and favorable pharmacokinetic properties, may reflect immune-associated metabolic alterations ^55^. In particular, lysoPE(18:3) has been reported to be more abundant in LuCa tissue ^49^. Serum levels of this phospholipid were significantly different in patients with cholangiocarcinoma (CCA), making it a potential biomarker to distinguish between early and late disease recurrence ^49,56^. 4-Hydroxybenzoic acid is a phenolic compound derived from dietary and microbiota metabolism, with a reported anti-cancer bioactivity ^57^.

Octadecylamine is a long-chain amine often reported in untargeted metabolomics studies as exogenous or derived from lipid or amino chemistry. However, this compound is also experimentally a common component of nanoparticles and drug-delivery constructs. For instance, nanoparticles made with octadecylamine are utilized as carriers for biocompounds in gene delivery and immunotherapy during cancer treatment that also include ovarian cancer ^58^.

In the ROC analysis of late-stage LuCa vs. late-stage CRC, we determined a series of metabolites with a combined AUC of 0.89 including metabolites described above, such as phenol sulfate, benzoic acid, 4-hydroxybenzoic acid, and sphingosineFor the dipeptide Ile-Ile it has been suggested that genetic variations related to isoleucine metabolism may be linked to a higher risk of LuCa. Specifically, a meta-analysis demonstrated that individuals carrying at least one valine allele (Ile/Val or Val/Val genotypes) had a statistically significant association with LuCa compared to those with the Ile/Ile genotype ^60^. Uracil is a pyrimidine base and RNA building block. Its metabolism is frequently altered in tumors, leading to accumulation that is a suitable candidate biomarker as part of the pyrimidine metabolism ^61^. It is clinically relevant for drug sensitivity, with uracil-DNA glycosylase (UNG) linked to LuCa response and has been detected in various tumor tissues including CRC ^62^. Uridine, a pyrimidine nucleoside essential for RNA synthesis and salvage pathways, can act as a metabolic fuel under glucose restriction, and its related enzymes (UPP1, UCK2) have been implicated in tumor biology as potential biomarkers and therapeutic targets ^63,64^. Lastly, linoleylcarnitine has repeatedly been linked to tumor progression and can impact oxidative stress regulation and energy adaptation ^59^.

We have also performed ROC analysis for OvCa at the early versus late stages. Although the AUC values in this case were moderate, these stage-associated metabolites can provide mechanistic insights into disease progression. When combined with other biomarkers, these compounds have the potential to substantially improve diagnostic performance. Epipregnanolone sulfate is a neurosteroid derived from progesterone. This steroid is known to modulate receptors in the nervous system, such as the GABAA receptors. While studies on the direct effects of epipregnanolone sulfate on ovarian cancer is limited, its potential role can be related to the closely related compound allopregnanolone. Allopregnanolone has been demonstrated to significantly impact ovarian physiology by modulating processes like apoptosis and angiogenesis, which are crucial for cancer development. It is therefore hypothesized that epipregnanolone sulfate could similarly impact ovarian cancer development and progression through these neuroactive steroid pathways ^65,66^. A predominant compound class that was significantly altered are acylcarnitines that were also observed in the late-stage LuCa vs. late-stage CRC analysis. Tetradecanoylcarnitine, palmitoylcarnitine, and linoleylcarnitine are involved in fatty acid metabolism for mitochondrial transport and oxidation ^67^. Altered acylcarnitine profiles have been previously reported in colorectal, lung, and ovarian cancer and specifically palmitoylcarnitine was changed in colorectal cancer cells ^68,69^.

The high discriminatory performance across all reported univariate and ROC comparisons in this study highlights the potential of metabolomics as a diagnostic tool. Particularly in settings where tissue access is limited due to the invasive sample collection or conventional imaging provides ambiguous results. The identified metabolite panels could thus serve as the basis for developing non-surgical diagnostic assays, such as blood-based tests (liquid biopsies), to enhance early cancer detection and guide clinical decisions.

In this study, we performed untargeted mass spectrometry-based metabolomics analysis to investigate a human plasma cohort of lung, colorectal and ovarian cancer patients. Our analysis revealed 3-hydroxypyridine sulfate, *S*-propenylcysteine, benzoic acid, lysoPC species, sphingosine, and several acylcarnitines as part of distinct early and late-stage signatures. Shared features across cancers suggest common metabolic stress and inflammation pathways, while cancer-specific metabolites reflect tissue-specific biological mechanisms and in some cases, diet or microbiome derived exposures. Notably, xenobiotic conjugates such as phenol sulfate and 3-hydroxypyridine sulfate highlight the interconnected influence of host metabolism and environmental or microbial factors, whereas lipids and redox-active species align with already established cancer mechanisms. Combination of the metabolite panels yielded excellent AUC values of up to 0.95 in the ROC curve analysis for differentiating both between cancer types and stage progression, with early-stage discrimination highlighting the potential utility in detection before clinical progression. Our findings expand on the number of candidate blood-based cancer biomarkers and additionally provide a biological framework for integrating metabolomics with microbiome, and genomic data in future validation studies.

## Methods

### Ethical approval

The study was approved by the Swedish Ethical Review Authority (EPM dnr 2019-00222). The research was in line with informed consents obtained from patients included in U-CAN (EPN Uppsala 2010-198).

### Study design

Plasma samples were obtained through the U-CAN project. The plasma samples (n = 710) were randomly divided into three analytical batches, A (n = 229), B (n = 239) and C (n = 242) plasma samples. Details are reported in Table 1. Each batch included a pooled quality control (QC) sample to monitor the analytical performance across the UHPLC–MS/MS analysis.

### Metabolite extraction

Plasma samples were kept on ice throughout the extraction procedure. The standard extraction protocol used an aliquot of 50 µL of plasma per sample. To each sample, 10 μL of internal standard mixture (5 μg/mL of tyrosine-^13^C_9_,^15^N, 10 μg/mL of phenylalanine-^13^C_9_,^15^N, and 20 μg/mL of valine-^13^C_5_) and 240 μL of ice-cold methanol were added. Prior to extraction, pooled quality control (QC) samples were prepared by combining 5 µL aliquots from each sample. QC samples underwent the same extraction procedure as the individual samples. Following methanol addition, all samples were vortexed briefly and stored at -20 °C for 1 h to precipitate proteins. Samples were then centrifuged at 14,500 rpm for 5 min and 4 °C and the supernatant was transferred to clean Eppendorf tubes. The supernatants were freeze-dried (Labconco, FreeZone 4.5 L Benchtop Freeze-Dryer) and the dried extracts were reconstituted in 50 µL of 5% acetonitrile in water, transferred to LC–MS vials, and analyzed by UHPLC–MS/MS. For samples with <50 µL available, the plasma volume was recorded, and the volumes of internal standard, methanol and reconstitution solution were adjusted proportionally to maintain the same extraction ratio.

### UHPLC–MS/MS analysis

The UHPLC–MS/MS analysis was performed in a Maxis II ETD Q-TOF mass spectrometer (Bruker Daltonics, Germany) using ESI as the source with an Elute UHPLC (Bruker Daltonics, Germany). The separation was performed with an Acquity UPLC HSS T3 column (1.8 μm, 100 × 2.1 mm) from Waters Corporation. For mobile phases, A contained milli-Q water with 0.1% formic acid and B contained LC−MS grade methanol with 0.1% formic acid. The temperature of the column and the autosampler were kept at 40 °C and at 4 °C, respectively. The flow rate was set to 0.20 mL/min with an injection volume of 5 μL. The gradient used was as follows: 0–2 min, 0% B; 2–15 min, 0–100% B; 15–16 min, 100% B; 16–17 min, 100-0% B; 17–21 min, 0% B. The system was controlled using the Compass HyStar software package from Bruker (Bruker Daltonics, Germany). The MS acquisition was performed in positive and negative ionization modes. The mass range was set from *m/z* = 50–1,200. Data acquisition was performed in AutoMSMS mode (data dependent acquisition, DDA) with a cycle time of 0.5 s and a ramped collision energy from 20 to 50 eV. A solution of sodium formate [10 mM in a mixture of 2-propanol/water (1/1, v/v)] was used for internal calibration at the beginning of each LC–MS experiment in a segment between 0.10 and 0.31 min.

### Statistical information

The LC−MS raw data were converted into mzML format using MS convert (ProteoWizard) and were further processed with the XCMS package in R (version 4.3.2). Feature tables were filtered to remove signals <1 min and >18 min and low intensity features (average ion count <10,000). First, for each batch independently, we applied SERRF (Systematic Error Removal using Random Forest), a QC-based method, to correct for within-batch variations that may arise from sample preparation or instrumental drift^36^. Univariate analysis was carried out in MetaboAnalyst 6.0 and GraphPad Prism 9.0 (GraphPad Inc., San Diego, USA) using unpaired t-tests (two-tailed) or one-way ANOVA with Tukey’s HSD post-hoc analysis when comparing multiple groups ^70^. P < 0.05 were considered significant, and box plots were generated for visualization in GraphPad. Multivariate analysis (PCA) was performed in MetaboAnalyst and R (version 4.4.3) with pareto scaling.

For combining the three analytical batches, the batch correction algorithm ComBat in MetaboAnalyst was used ^37^. ROC curve analysis was performed in Python (v3.9) using scikit-learn to assess the discriminatory performance of selected metabolites between two study groups. Binary class labels were assigned (0/1) and individual metabolite performance was assessed using logistic regression (solver = “liblinear”), with the area under the ROC curve (AUC) used as the performance metric. The top 20 metabolites ranked by individual AUC were selected for subsequent combinatorial analysis. All possible 10-metabolite combinations were evaluated using logistic regression models, and the combination achieving the highest AUC was selected for further consideration. To ensure biological relevance, candidate features were subsequently curated based on metabolite annotation, and known exogenous compounds (e.g., drugs) were excluded from the final panel. ROC curves for the curated metabolite panel were then generated, and 95% confidence intervals were estimated using nonparametric bootstrap resampling (n = 1000 iterations). The combinatorial analysis was performed in an exploratory manner to identify high-performing candidate biomarker panels.

Metabolite annotation was performed using MS/MS fragmentation experiments (DDA, 20–50 eV), and spectra matching with our in-house spectral libraries, online public databases (HMDB and MoNA) and computational tools like SIRIUS to increase the confidence of metabolite identification ^71,72^. The annotation confidence levels (CLs) were assigned according to the Schymanski et al. criteria, and the metabolites used in biological interpretation were restricted to those reaching at least CL 2; annotated based on spectral similarity with online databases or computational fragmentation tools (Table S1, Figure S9) ^73^.

## Supporting information

Tsiara_Supporting Information

## Data Availability

All data produced in the present study are available upon reasonable request to the authors

## Acknowledgements

This study was funded by the Swedish Research Council (D.G. 2020-04707) and the Swedish Cancer Society (D.G. 22 2449 Pj, D.G. 25 4898 Pj, T.S, 211719Pj, T.S. 24 3831 Pj). We are grateful to U-CAN for access to plasma samples from Uppsala Biobank with support from the Swedish Government (SRA grant CancerUU),

## Author Contribution Statement

IT: methodology, investigation, formal analysis, data curation, validation, visualization, writing – original draft. EV: investigation, validation, writing – original draft. JE, IS: methodology, formal analysis, data curation. NR., FY, AJ, AIA: methodology. TS: supervision, project administration, funding acquisition, data curation, conceptualization, writing – review & editing. DG: supervision, project administration, funding acquisition, methodology, data curation, conceptualization, writing – original draft.

## Ethics declarations

The authors declare no competing interests for this study.

