## Supplementary material for "Distinct Metabolic Signatures Distinguish Lung, Colorectal and Ovarian Cancer": Tsiara_Supporting Information

### Figures

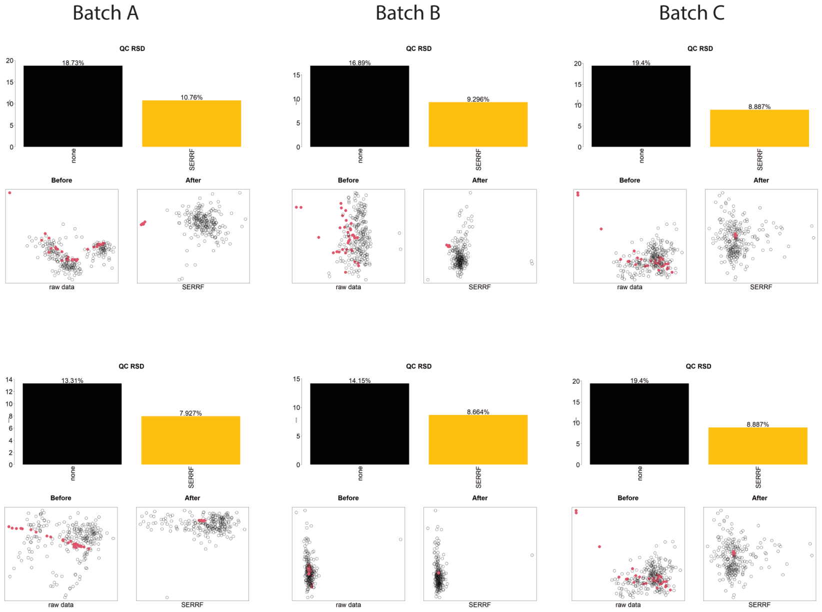


**Figure S1.** SERRF normalization results. Positive ionization mode (top) and negative ionization mode (bottom).

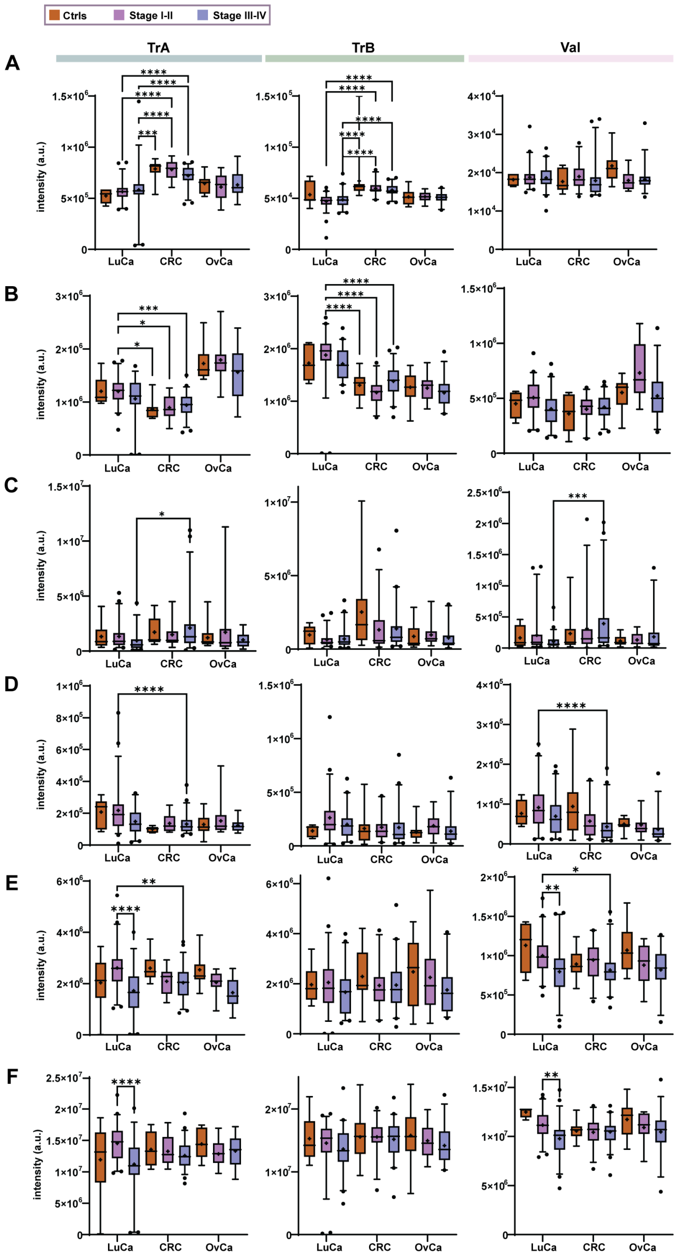


**Figure S1.** Statistically significant metabolites found in two independent batches (one-way ANOVA with Tukey's HSD post-hoc analysis, ****$p \leq 0.0001$; ***$p \leq 0.001$; **$p \leq 0.01$; *$p \leq 0.05$). A) Benzoic acid, B) *S*-propenylcysteine, C) phenol sulfate, D) cyclo(Leu-Pro), E) lysoPC (14:0) and F) lysoPC (18:2).

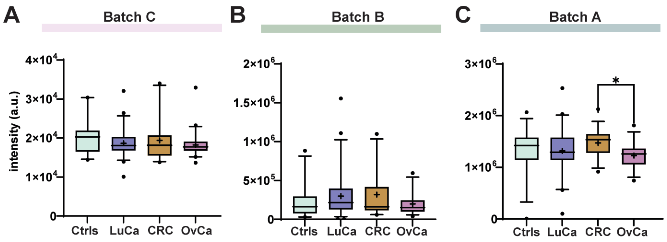


**Figure S3.** Batches that did not show consistent alteration in the sex-dependent investigation of females. A) Benzoic acid, B) sphingosine and C) lysoPC (20:3).

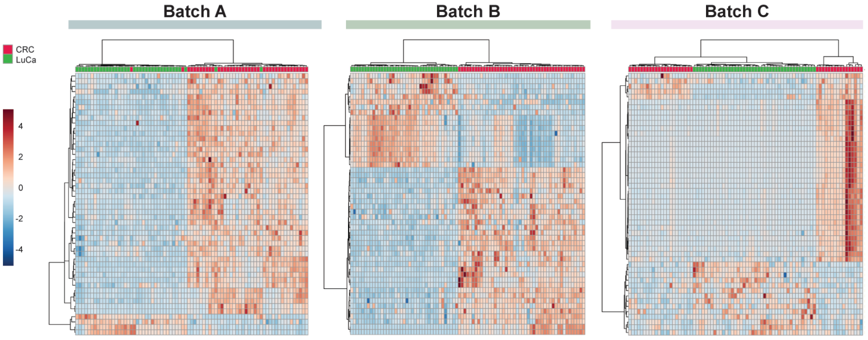


**Figure S4.** Heatmaps of male individuals showing the top 50 significantly altered metabolite features (unpaired t-test, $p < 0.05$) between LuCa and CRC in positive ionization mode.

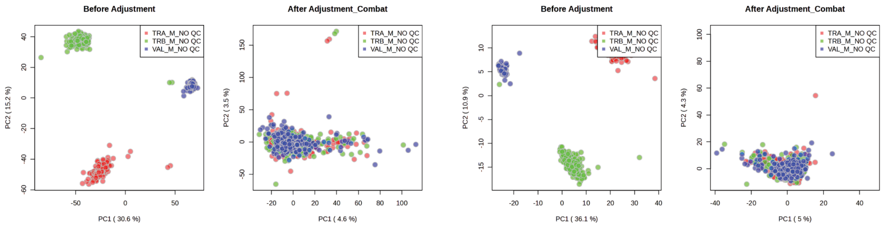


**Figure S5**. Combat algorithm results for batch normalization. Positive ionization mode (left) and negative ionization mode (right).

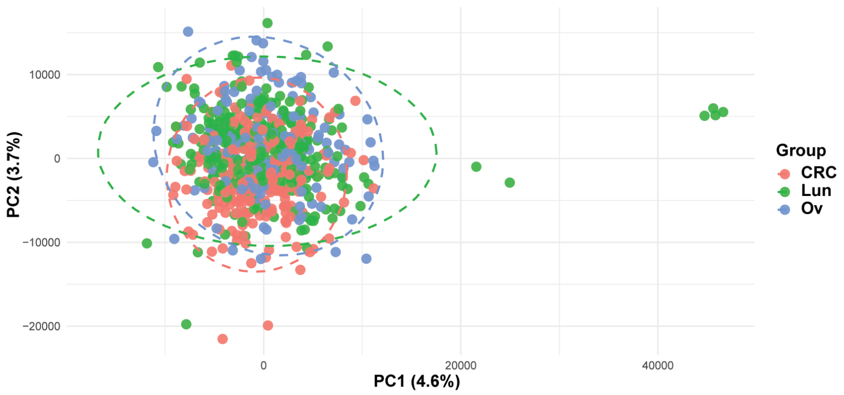


**Figure S6.** PCA scores plot of the merged dataset with the ComBat algorithm for all three cancer groups in positive ionization mode.

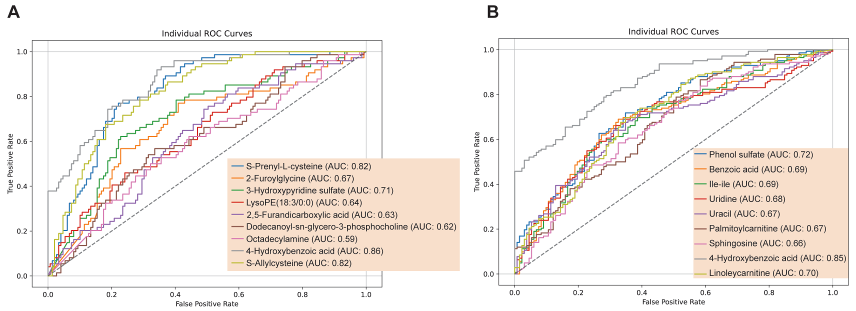


**Figure S7.** Individual ROC curves for A) early-stage and B) late-stage comparison of LuCa and CRC.

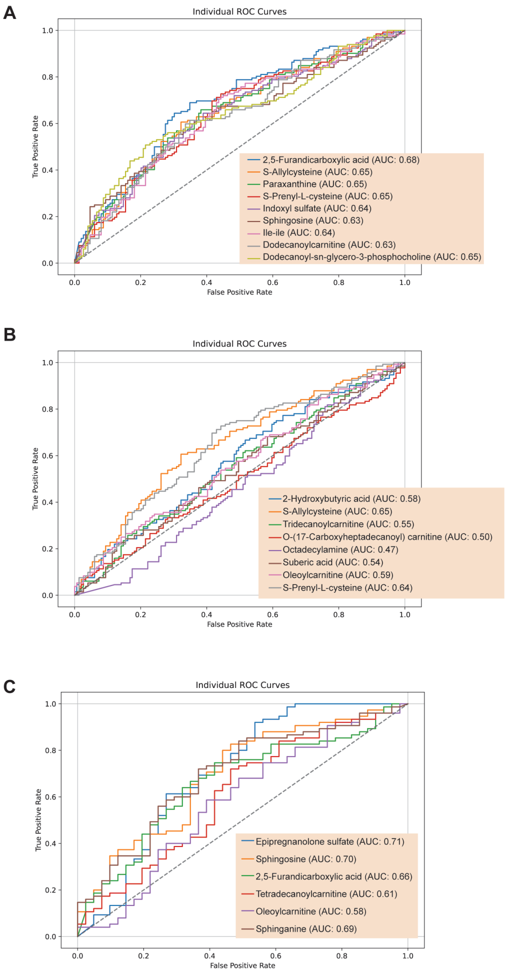


**Figure S8.** Individual ROC curves for early-stage vs late-stage A) LuCa B) CRC and C) OvCa.

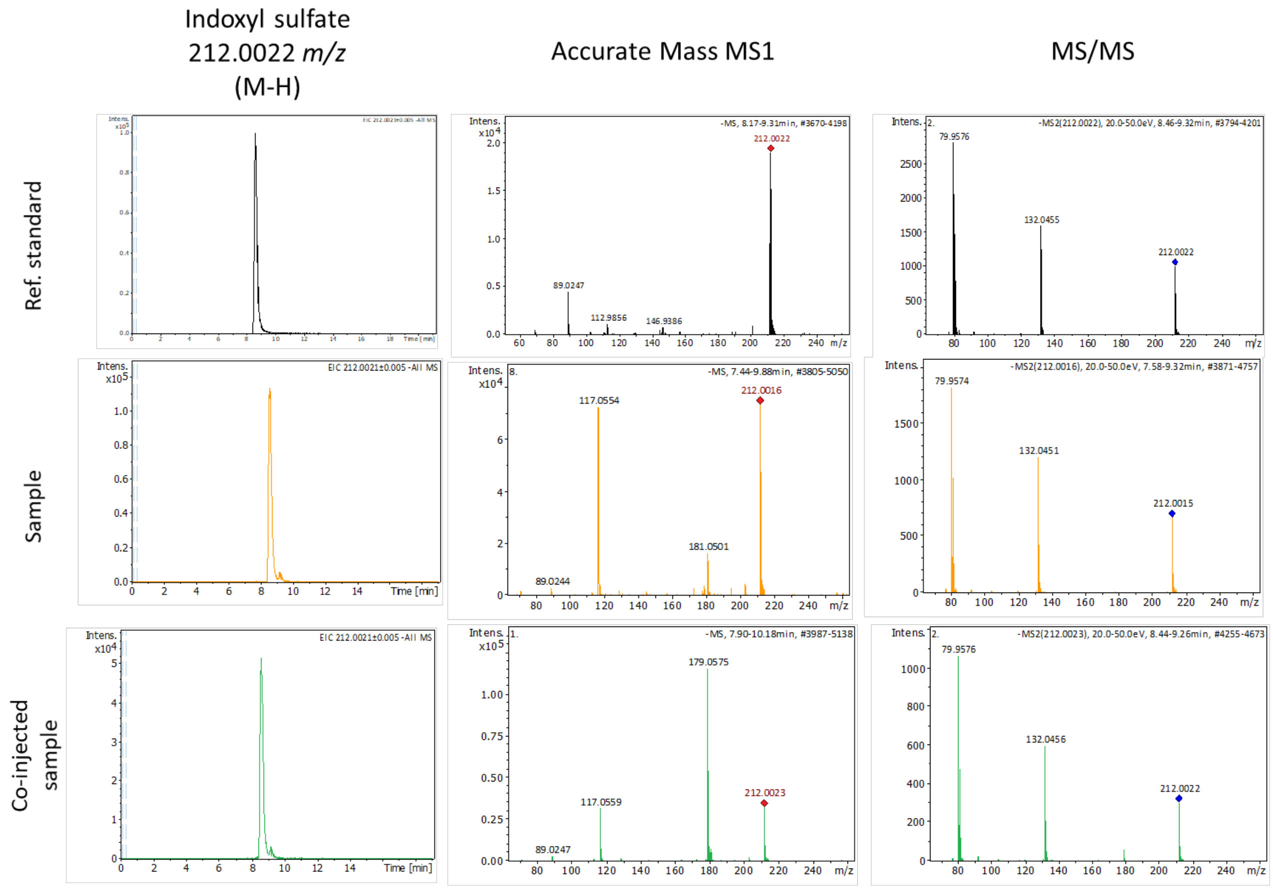


**Figure S9.** General workflow for metabolite identification. Metabolites were identified in reference samples based on co-elution with authentic reference standards and matching accurate-mass MS1 and MS/MS spectra.

#### Tables

**Table S1.** All validated metabolites with annotated confidence levels (CL) in the respective mode (Adduct).

*CL1: Validation with authentic synthetic or commercial standards; CL 2: Metabolite structure validation based on MS2 spectra with experimental spectra from literature or library sources, or fragmentation pattern comparison using computational tools; CL3: Tentative structure validation based on MS2 experimental spectra.*

| # | Monoisotopic mass (Da) | Compound name | ID | Adduct | CL |
|---|---|---|---|---|---|
| 1 | 104.0475 | 2-Hydroxybutyric acid | HMDB0000008/ HMDB0000011 | $[M - H]^-$ | 2 |
| 2 | 108.0577 | *p*-Cresol | HMDB0001858 | $[M - H]^-$ | 2 |
| 3 | 112.0274 | Uracil | HMDB0000300 | $[M + H]^+$ | 2 |
| 4 | 113.0841 | Epsilon-caprolactam | HMDB0062769 | $[M + H]^+$ | 2 |
| 5 | 118.0632 | 2-Hydroxyisovaleric acid/3-Hydroxyvaleric acid | HMDB0000407/ HMDB0000531 | $[M - H]^-/[M + Na]^+$ | 2 |
| 6 | 122.0368 | Benzoic acid | HMDB0001870 | $[M + H]^+$ | 1 |
| 7 | 128.0627 | Naphthalene | HMDB0029751 | $[M + H]^+$ | 2 |
| 8 | 131.0947 | L-Norleucine | HMDB0001645 | $[M + H]^+$ | 2 |
| 9 | 136.0374 | Erythronic acid/Threonic acid | HMDB0000613/ HMDB0000943 | $[M - H]^-$ | 2 |
| 10 | 138.0317 | 4-Hydroxybenzoic acid | HMDB0000500 | $[M + H]^+$ | 2 |
| 11 | 150.0531 | D-Arabinose | HMDB0029942 | $[M - H]^-$ | 2 |
| 12 | 152.0476 | Phenoxyacetic acid/Methylparaben/Methyl 2-hydroxybenzoate | HMDB0031609/ HMDB0032572/ HMDB0034172 | $[M - H]^-/[M + H]^+$ | 3 |
| 13 | 156.0065 | 2,5-Furandicarboxylic acid | HMDB0004812 | $[M + H]^+$ | 2 |
| 14 | 161.0512 | *S*-Propenylcysteine | HMDB0302212 | $[M + H]^+$ | 2 |
| 15 | 169.0378 | 2-Furoylglycine | HMDB0000439 | $[M - H]^-$ | 2 |
| 16 | 172.1102 | 8-Oxononanoic acid | HMDB0341050 | $[M - H]^-$ | 2 |
| 17 | 173.9989 | Phenol sulfate | HMDB0060015 | $[M - H]^-$ | 1 |
| 18 | 174.0894 | Suberic acid | HMDB0000893 | $[M - H]^-$ | 2 |
| 19 | 174.1118 | Arginine | HMDB0000517 | $[M + H]^+$ | 2 |
| 20 | 174.1259 | 3-Hydroxynonanoic acid | HMDB0031513 | $[M - H]^-$ | 2 |
| 21 | 174.9940 | 3-hydroxypyridine sulfate | HMDB0240652 | $[M - H]^-$ | 1 |
| 22 | 176.0546 | Allantoic acid | HMDB0001209 | $[M + Na]^+$ | 2 |
| 23 | 176.0952 | Cotine | HMDB0001046 | $[M + H]^+$ | 2 |
| 24 | 180.0636 | Fructose/Sorbose/ Tagatose | HMDB0000660/ HMDB0001266/ HMDB0003418 | $[M - H]^-$ | 2 |
| 25 | 180.0644 | Paraxanthine | HMDB0001860 | $[M - H]^-/[M + Na]^+$ | 2 |
| 27 | 180.0644 | Theophylline | HMDB0001889 | $[M - H]^-$ | 1 |
| 28 | 181.0741 | L-Tyrosine | HMDB0000158 | $[M - H]^-$ | 2 |
| 29 | 182.9993 | Saccharin | HMDB0029723 | $[M - H]^-$ | 2 |
| 30 | 188.0144 | *p*-Cresol sulfate | HMDB0011635 | $[M - H]^-$ | 1 |
| 31 | 188.1412 | 9-Hydroxydecanoic acid | HMDB0033201 | $[M + Na]^+$ | 2 |
| 32 | 188.1415 | 10-Hydroxydecanoic acid | HMDB0244272 | $[M - H]^-$ | 2 |

| | | | | | |
|---|---|---|---|---|---|
| 33 | 189.0825 | *S*-Prenyl-L-cysteine | HMDB0012286 | $[M + H]^+$ | 2 |
| 34 | 189.9938 | Resorcinol sulfate | HMDB0304905 | $[M - H]^-$ | 1 |
| 35 | 192.0271 | Citric acid | HMDB0000094 | $[M - H]^-$ | 1 |
| 36 | 192.0637 | Quinic acid | HMDB0003072 | $[M - H]^-$ | 2 |
| 37 | 196.1214 | Cyclo(L-prolyl-L-valyl) | HMDB0240493 | $[M + H]^+$ | 2 |
| 38 | 202.1207 | Sebacic acid | HMDB0000792 | $[M - H]^-$ | 2 |
| 39 | 203.1157 | L-Acetylcarnitine | HMDB0000201 | $[M + H]^+$ | 2 |
| 40 | 210.1369 | Cyclo(Leu-Pro) | HMDB0034276 | $[M + H]^+$ | 1 |
| 41 | 213.0095 | Indoxyl sulfate | HMDB0000682 | $[M - H]^-$ | 1 |
| 42 | 214.1572 | 11-Oxododecanoic acid | CID547024 | $[M - H]^-$ | 2 |
| 43 | 214.1572 | 5-Oxododecanoic acid | CID3250790 | $[M - H]^-$ | 2 |
| 44 | 216.1364 | Undecanedioic acid | HMDB0000888 | $[M - H]^-$ | 2 |
| 45 | 216.1728 | 3-Hydroxydodecanoic acid | HMDB0000387 | $[M - H]^-$/$[M + Na]^+$ | 2 |
| 46 | 231.0203 | Paracetamol sulfate | HMDB0059911 | $[M - H]^-$ | 2 |
| 47 | 240.0998 | 3-Carboxy-4-methyl-5-propyl-2-furanpropionic acid | HMDB0061112 | $[M + H]^+$/$[M - H]^-$ | 2 |
| 48 | 242.1885 | 6-Ketomyristic acid | HMDB0030982 | $[M - H]^-$ | 2 |
| 49 | 244.0698 | Uridine | HMDB0000296 | $[M - H]^-$ | 2 |
| 50 | 244.1790 | Isoleucyl-Isoleucine | HMDB0028910 | $[M + H]^+$ | 2 |
| 51 | 244.2041 | 2-Hydroxymyristate/ 3-Hydroxymyristic acid | HMDB0002261/ HMDB0010731 | $[M - H]^-$ | 2 |
| 52 | 250.1208 | 3,4-Dihydro-6-hydroxy-2,5,7,8-tetramethyl-2H-1-benzopyran-2-carboxylic acid | HMDB0038804 | $[M - H]^-$ | 2 |
| 53 | 250.1570 | Hexyl benzoate | HMDB0040431 | $[M - H]^-$ | 2 |
| 54 | 258.1834 | Tetradecanedioic acid | HMDB0000872 | $[M - H]^-$ | 2 |
| 55 | 261.0308 | Tyrosine sulfate | CID514186 | $[M - H]^-$ | 2 |
| 56 | 262.0820 | Glutamylaspartic acid | HMDB0028815 | $[M - H]^-$ | 2 |
| 57 | 262.1167 | Nopalinic acid | HMDB0029437 | $[M + H]^+$ | 2 |
| 58 | 269.3083 | Octadecylamine | HMDB0029586 | $[M + H]^+$ | 2 |
| 59 | 271.2514 | Hexadecasphingosine | HMDB0242181 | [M + H]+ | 2 |
| 60 | 271.2514 | *N*-(2-hydroxyethyl)tetradecanamide | CID8890 | $[M + H]^+$ | 2 |
| 61 | 272.1990 | Pentadecanedioic acid | HMDB0340876 | $[M - H]^-$ | 2 |
| 62 | 278.1886 | 2-Phenoxyundecanoic acid | CID22069708 | $[M - H]^-$ | 2 |
| 63 | 281.2719 | Oleamide | HMDB0002117 | $[M + Na]^+$ | 2 |
| 64 | 285.1392 | Piperine | HMDB0029377 | $[M + H]^+$ | 2 |
| 65 | 286.2147 | Hexadecanedioic acid | HMDB0000672 | $[M - H]^-$ | 2 |
| 66 | 292.0906 | Edetic Acid | HMDB0015109 | $[M - H]^-$ | 2 |
| 67 | 299.2825 | Sphingosine | HMDB0000252 | $[M + H]^+$ | 1 |
| 68 | 301.2982 | Sphinganine | HMDB0000269 | $[M + H]^+$ | 2 |
| 69 | 303.2049 | 3-Hydroxyoctanoyl carnitine | HMDB0061634 | $[M + H]^+$ | 2 |
| 70 | 306.1065 | Edetic Acid methyl ester | CID17826827 | $[M - H]^-$/$[M + H]^+$ | 2 |

| | | | | | |
|---|---|---|---|---|---|
| 71 | 327.0955 | Acetaminophen glucuronide | HMDB0010316 | [M - H]- | 2 |
| 72 | 331.2362 | 3-hydroxydecanoyl carnitine | HMDB0061636 | [M + H]+ | 2 |
| 73 | 344.2795 | Dodecanoylcarnitine | HMDB0002250 | [M + H]+ | 2 |
| 74 | 357.2880 | Tridecanoylcarnitine | HMDB0241308 | [M + H]+ | 2 |
| 75 | 370.1816 | Androsterone sulfate/Epiandrosterone sulfate | HMDB0002759/ HMDB0062657 | [M - H]- | 2 |
| 76 | 372.3108 | Tetradecanoylcarnitine | HMDB0005066 | [M + H]+ | 2 |
| 77 | 387.2988 | 3-Hydroxytetradecanoyl carnitine | HMDB0061640 | [M + H]+ | 2 |
| 78 | 391.2728 | N-Arachidonoyl Serine | HMDB0242003 | [M + H]+ | 2 |
| 79 | 398.2129 | Epipregnanolone sulfate | CID73755084 | [M - H]- | 2 |
| 80 | 400.3421 | Palmitoylcarnitine | HMDB0000222 | [M + H]+ | 2 |
| 81 | 424.3421 | Linoleyl carnitine | HMDB0006469 | [M + H]+ | 2 |
| 82 | 426.3577 | Oleoylcarnitine | HMDB0005065 | [M + H]+ | 2 |
| 83 | 439.2702 | Dodecanoyl-sn-glycero-3-phosphocholine | HMDB0240736 | [M + H]+ | 2 |
| 84 | 457.3409 | O-(17-Carboxyheptadecanoyl)carnitine | HMDB0240777 | [M + Na]+ | 2 |
| 85 | 475.2706 | LysoPE(18:3) | HMDB0011508 | [M + H]+ | 2 |
| 86 | 477.2859 | LysoPE(18:2) | HMDB0011507 | [M + H]+ | 2 |
| 87 | 479.3017 | LysoPE(18:1) | HMDB0011505 | [M + H]+ | 2 |
| 88 | 481.3098 | LysoPC(15:0) | HMDB0010381 | [M + H]+ | 2 |
| 89 | 495.3329 | LysoPC(16:0) | HMDB0010382 | [M + H]+ | 2 |
| 90 | 503.3012 | LysoPC(20:3) | HMDB0011515 | [M + Na]+ | 2 |
| 91 | 523.3638 | LysoPC(18:0) | HMDB0010384 | M + H]+/[M + Na]+ | 2 |
| 92 | 542.2734 | Cortolone-3-glucuronide | HMDB0010320 | [M - H]- | 2 |
| 93 | 776.6862 | Thyroxine/ Dextrothyroxine | HMDB0000248/ HMDB0014651 | [M + H]+ | 2 |

**Table S2**. Top 20 metabolites with the greatest ability to distinguish between early stages and late stages of LuCa and CRC, ranked by absolute ROC AUC (max(AUC, 1 − AUC)).

| **Early-stage LuCa vs CRC** | | | |
|---|---|---|---|
| **Metabolite** | **Raw AUC** | **Discriminative AUC** | **Upregulated Group** |
| Unknown | 0.932 | 0.932 | CRC I–II |
| 4-Hydroxybenzoic acid | 0.865 | 0.865 | CRC I–II |
| S-Prenyl-L-cysteine | 0.823 | 0.823 | LuCa I–II |
| S-Propenylcysteine | 0.819 | 0.819 | LuCa I–II |
| Benzoic acid | 0.202 | 0.798 | CRC I–II |
| 3-hydroxypyridine sulfate | 0.713 | 0.713 | LuCa I–II |
| Resorcinol sulfate | 0.702 | 0.702 | LuCa I–II |
| 2-Furoylglycine | 0.672 | 0.672 | LuCa I–II |
| Unknown | 0.329 | 0.671 | CRC I–II |
| Methyl 2-hydroxybenzoate | 0.329 | 0.671 | CRC I–II |
| Cyclo(Leu-Pro) | 0.663 | 0.663 | LuCa I–II |
| LysoPC(20:3) | 0.339 | 0.661 | CRC I–II |

| | | | |
|---|---|---|---|
| Quinic acid | 0.659 | 0.659 | LuCa I–II |
| Erythronic acid/Threonic acid | 0.349 | 0.651 | CRC I–II |
| Naphthalene | 0.645 | 0.645 | LuCa I–II |
| 5-oxododecanoic acid | 0.355 | 0.645 | CRC I–II |
| LysoPE(18:3) | 0.644 | 0.644 | LuCa I–II |
| Uridine | 0.359 | 0.641 | CRC I–II |
| LysoPE(18:1) | 0.636 | 0.636 | LuCa I–II |
| L-Tyrosine | 0.633 | 0.633 | LuCa I–II |
| **Late-stage LuCa vs CRC** | | | |
| **Metabolite** | **Raw AUC** | **Discriminative AUC** | **Upregulated Group** |
| Unknown | 0.926 | 0.926 | CRC III–IV |
| 4-Hydroxybenzoic acid | 0.85 | 0.85 | CRC III–IV |
| Phenol sulfate | 0.724 | 0.724 | CRC III–IV |
| Linoleyl carnitine | 0.703 | 0.703 | CRC III–IV |
| 3-Hydroxydodecanoic acid | 0.695 | 0.695 | CRC III–IV |
| 3-Carboxy-4-methyl-5-propyl-2-furanpropionic acid | 0.695 | 0.695 | CRC III–IV |
| Benzoic acid | 0.694 | 0.694 | CRC III–IV |
| Methyl 2-hydroxybenzoate | 0.688 | 0.688 | CRC III–IV |
| Isoleucyl-Isoleucine | 0.687 | 0.687 | CRC III–IV |
| Glutamylaspartic acid | 0.682 | 0.682 | CRC III–IV |
| Uridine | 0.68 | 0.68 | CRC_III–IV |
| 3-Hydroxyvaleric acid | 0.678 | 0.678 | CRC III–IV |
| Tridecanoylcarnitine | 0.677 | 0.677 | CRC III–IV |
| Uracil | 0.675 | 0.675 | CRC III–IV |
| Palmitoylcarnitine | 0.672 | 0.672 | CRC III–IV |
| LysoPC(20:3) | 0.668 | 0.668 | CRC III–IV |
| Erythronic acid/Threonic acid | 0.667 | 0.667 | CRC III–IV |
| 8-oxononanoic acid | 0.662 | 0.662 | CRC III–IV |
| Naphthalene | 0.661 | 0.661 | LuCa III–IV |
| Paracetamol sulfate | 0.657 | 0.657 | LuCa III–IV |
